# Epidemiological analysis of the gender-age structure of hospitalized patients with COVID-19 and their mortality in 2020-2021

**DOI:** 10.1101/2023.02.09.23285515

**Authors:** Evgeny M. Voronin, Izabella A. Khrapunova, Andrey S. Pechenik, Olga A. Kravtsova, Zhang Chen, Mikhail P. Kostinov, Marina N. Loktionova, Irina V. Yakovleva, Vasily G. Akimkin, Victoria A. Brazhnik

## Abstract

**Background:** The pandemic of the new coronavirus infection COVID-19 determines the relevance of conducting a study of the gender-age structure of hospitalized and deceased among the population of Moscow on the example of one of the city clinical hospitals in 2020 – 2021.

The aim of the work is to identify the patterns of the epidemic process of COVID-19 in connection with the gender and age characteristics of hospitalized adults and to establish the sex and age groups most susceptible to hospitalization and mortality from COVID-19.

**Materials and methods:** The analysis of the structure of hospitalized and deceased from COVID-19 in the context of their gender and age composition was carried out. The data of the statistical accounting form F-60u/lech “Journal of infectious diseases” of adults hospitalized in one of the city clinical hospitals of Moscow in 2020 – 2021 were used. Data processing was carried out by a set of standard statistical methods. To identify the true risk groups for hospitalization and mortality, a statistical correction of the sex and age composition of the population of Moscow was used.

**Results and discussion:** Using standard statistical methods in combination with the use of statistical correction of sex and age composition, data on the true risk groups for hospitalization and mortality among the population of Moscow in 2020 – 2021 were obtained.

**Conclusion:** The results obtained in our independent study on the true risk groups for hospitalization and mortality among the population of Moscow complement and introduce new knowledge about the true risk groups for hospitalization and mortality in COVID-19. The patterns identified in this epidemiological analysis are an important component of epidemiological surveillance for making managerial decisions to prevent the spread of SARS-CoV-2 and planning for the provision of inpatient medical care to established gender and age risk groups.

## Introduction

COVID-19 is a dangerous disease that can occur as a mild to severe acute respiratory viral infection. The SARS-CoV-2 virus can infect different organs by direct infection or through the body’s immune response [1-4]. The start of the pandemic has been linked to an outbreak of SARS-CoV-2 infection in Wuhan, China, in December 2019, after which the coronavirus rapidly spread worldwide [5]. Recent epidemiological studies show that coronavirus infection with COVID-19 is associated with a high rate of hospitalization in intensive care units [6]. The lethality of COVID-19 has not been accurately established, with literature suggesting a range of 3-6%, but some researchers have suggested that it may be severely underestimated for political reasons [7].

The incidence of the new coronavirus infection COVID-19 in the world, Russia and Moscow is now rapidly declining. According to the Internet resource, the highest incidence rate in the world was recorded on 19.01.2022, when the number of people falling ill per day was 4,079,835. As of June 2022, the incidence is represented by the lowest number of cases. Thus, on 05.06.2022, the number of reported cases worldwide was 254,040. We have seen a sharp decline in the incidence of COVID-19 in 4.5 months by a factor of about 16: from 4 million to 0.25 million.

In Russia, the number of COVID-19 cases in February (11.02.2022) this year peaked at over 200,000 (203,949). As of 05.06.2022, the number of people falling ill was less than 4,000 (3,786); for 4 months the incidence has decreased more than 50 times. In Moscow, the maximum number of people sickened per day was also recorded in February (03.02.2022) this year - about 27 thousand people (26904), on 05.06.22 the minimum number was recorded - 185 people sickened per day, which is 141 times lower than the maximum values [8,9].

The end of the pandemic was a collaborative effort between scientists, governments and health authorities around the world, who imposed quarantine measures, developed effective vaccines rapidly and succeeded in mass vaccination of the public, including in economically developing countries [10-14]. The COVID-19 pandemic has a ripple effect and is accompanied in the course of the epidemic by a change in the pathogenic properties of the agent as a result of successive genetic modifications that result in successive changes in the dominant circulating strains of SARS-CoV-2 [15-18].

The time has come to take stock of the extraordinary epidemic situation that has claimed millions of lives. To avert the human, economic and social toll of the pandemic, a comprehensive multi-factorial retrospective epidemiological analysis, with elements of clinical epidemiology [19], is needed to assess the positive and negative aspects of the endgame, to draw objective conclusions and to suggest better, perhaps unconventional, solutions that might mitigate such events in the future.

One of the most obvious questions in such retrospective epidemiological analyses is whether and how morbidity and mortality are related to sex and age.

Therefore, to assess the significance of gender and age among hospitalized patients and their mortality from COVID-19 according to data published in open Russian and foreign scientific medical journals, we conducted a search of scientific papers on the relevant topic. A systematic search was performed in the PubMed database, the WHO COVID-19 database, and Web of Science. We also used, to a small extent, full-text original articles and two preprint servers.

We used the following keywords to generate the search queries: ‘COVID-19’, ‘SARS-CoV-2’, ‘hospitalization’, ‘mortality’, ‘sex’, ‘age’, ‘risk group’. For the search, we considered ‘systematic review’ or ‘meta-analysis’ publications with only adult patients (over 18 years of age) for the period January 2020-June 2022.

An example search query for the PubMed database is as follows: ““COVID-19”[Mesh] OR “SARS-CoV-2”[Mesh] OR “Covid 19” [tw] OR “SARS-CoV-2”[tw] OR “Novel Coronavirus” [tw] AND (“Hospital Mortality”[Mesh] OR “Hospitalization”[Mesh] OR death* [tw]) AND (sex [tw] or (male [tw] and female [tw])) AND (“Middle Aged”[MeSH] OR age [tw]) AND (statist* [tw] OR character* [tw]) AND (Meta-Analysis [pt] OR Systematic Review [pt]) NOT “COVID-19 Vaccines”[Mesh] “.

A total of 29 studies were selected for a more detailed review. A sufficient level of specificity, reflecting the focus of our study, was identified in 10 published papers.

A number of published studies from Italy, Denmark, India, China [20, 21, 22, 23, 24], based on retrospective cohort studies analyzing significant rates of hospitalization and mortality from COVID-19, along with the effect of comorbidity on course severity and mortality, show a definite trend in the influence of gender and age characteristics. For example, researchers note that older men and those with comorbidities are at higher risk of adverse clinical outcomes, including mortality. The likelihood of contracting SARS-CoV-2 depends on age category and gender. Women under 35 years of age have a higher chance of infection than men, but hospitalization rates increase in men as age increases (> 35 years) (22, 25). Mortality rates in older men are higher than in women, but lower than in younger men (26). According to the European Centre for Disease Prevention and Control [23], in the Nordic region (excluding Iceland), 72% of hospital admissions were male. In the region as a whole (with the exception of Finland), a higher proportion of deaths (54.7%) were male. In Finland, the percentage of deaths in women and men was approximately the same. A systematic review [24] of a total of 76 555 adult patients hospitalized with COVID-19 concluded from ten studies that there was a higher risk of mortality for men than for women, in eight of these studies the risk was statistically significant. The same paper concluded that male gender is a significant risk factor in increased mortality from COVID-19 in adult patients. This association may be explained by physiological differences between men and women: men tend to have lower serum IgG antibody production, lower CD4+ T-cell levels and lower circulating ACE2. At the same time, female gender has been found to play a significant role in reducing the risk of mortality from COVID-19.

A meta-analysis by Wei X. at all [27], performed on 39 studies with a total of 77,932 patients with COVID-19, concluded that men have a significantly higher risk of both severe course and mortality compared to women.

Quaresima V. at all [28] analyzed the hospitalization and mortality of patients with COVID-19 as a function of sex and age, using 367 women and 633 men in the province of Brescia (northern Italy). The results showed that the mean age at the time of hospitalization was similar for women and men, the overall mortality rate did not differ between the sexes, but the women who died were older. The authors concluded that the risk of death for hospitalized women and men was similar. This was the only paper we found in which the researchers pointed out that in the province of Brescia, with the same number of hospitalizations for women and men over 75 years old, the number of elderly women was about twice as high as the number of elderly men.

In an observational study of causes of death during the COVID-19 pandemic in India [29], age was identified as the strongest predictor of death, although in this respect COVID-19 is not fundamentally different from a significant number of other infectious diseases. There was also a significantly higher mortality rate in men after the first week of hospitalization.

Overall, the findings of a number of publications suggest that there is currently no single, unambiguously accepted position in defining the sex- and age-specific risk groups for hospitalization and mortality in COVID-19 disease. The majority of researchers estimate that characteristics such as older age, male gender and comorbidities are the most likely risk factors for mortality.

It should be noted that in none of the studies we reviewed did we find gender and age proportions in the population of hospitalized and deceased COVID-19 patients as an important and relevant criterion.

Thus, the aim of our study is to identify the patterns of the epidemic process of COVID-19 in relation to the gender and age characteristics of hospitalized adults and to identify the sex and age groups most susceptible to hospitalization and mortality from COVID-19. Answers to these questions can help to guide management decisions on prevention and control measures when similar epidemic situations arise in the future.

## Materials and methods

The study was conducted in accordance with the research plan of the Department of Epidemiology and Modern Methods of Vaccination, Faculty of Epidemiology, Sechenov First Moscow State Medical University, Ministry of Health of Russia (Sechenov University). The study was conducted in collaboration with the Scientific Group of Mathematical Methods and Epidemiological Forecasting of the Central Research Institute of Epidemiology of the Russian Federation and is a retrospective non-randomized epidemiological study.

The data for processing were obtained from the Form F-60u/Lech “Infectious Disease Logbook” of adults hospitalized in one of the city clinical hospitals in Moscow in 2020-2021. The analyzed sample size was 5629 hospitalized with COVID-19 over the period 2020-2021, of which 3106 (55.2%) were female and 2523 (44.8%) were male. The study sample was divided into age cohorts (people of the same year of birth), age groups (one group combining several age cohorts) according to WHO classification (young age, middle age, old age, old age and longevity) and by gender. Analysis of morbidity and mortality was carried out according to gender differences in different age cohorts and groups.

For comparative analysis of the gender structure of hospital admissions and mortality among them, a statistical correction of the sex and age composition (SCSAC) of the Moscow population according to the ratio of men to women in age cohorts was applied, based on the data of the Federal State Statistics Service (Rosstat) [30]. The calculated dimensionless coefficient of the SCSAC shows how many women there are per one man in each age group (and, if necessary, in each age cohort) in the sample under study.

Data on all indicators were prepared for further retrospective epidemiological analysis using Python (pandas package) [31, 32, 33]. For statistical data processing (graphing, approximation curves, determination of coefficients of determination (reliability of approximation), determination of percentages, etc.), the Excel statistical software package was used. Standard deviation was used to reflect the measure of dispersion of the mean. The confidence interval for the proportion by age of men and women among the diseased and in the general population in Moscow was calculated using binomial criterion. Differences in the distribution of the categorical variable “men/women” across age groups were examined using the chi-square criterion. To measure the strength of the association, Cramer’s measure V [34].

## Results and discussion

The COVID-19 pandemic has shed light on how important patient databases are for the development of knowledge about new diseases. Given the serious gaps in knowledge about COVID-19, they are of paramount importance for medical research.

The distribution of those hospitalized with COVID-19 by gender and age is shown in **Table 1** (baseline data).

**Table 1.** Distribution of those hospitalized with COVID-19 by gender and age group (years) in baseline data.

| WHO classification | Young age | Middle age | Older age | Senile age | Longevity |
| --- | --- | --- | --- | --- | --- |
| Age groups, years | 18-44 | 45-59 | 60-74 | 75-90 | 91+* |
| Number of age cohorts | 27 | 15 | 15 | 16 | 10 |
| Arithmetic mean of age in each group, years | $39,0 \pm 5,0$ | $53,1 \pm 4,3$ | $68,3 \pm 4,5$ | $83,1 \pm 3,9$ | $93,6 \pm 2,7$ |
| **Hospitalized** |  |  |  |  |  |
| Total number of hospitalized, abs. | 618 | 1140 | 2041 | 1661 | 169 |
| Proportion of age group among those hospitalized, % | 11,0 | 20,3 | 36,2 | 29,5 | 3,0 |
| Number of male hospitalized by age group, abs. | 356 | 602 | 941 | 578 | 46 |
| Number of female hospitalized by age group, abs. | 262 | 538 | 1100 | 1083 | 123 |
| Proportion of male hospitalized by age group, % | 57,6 | 52,8 | 46,1 | 34,8 | 27,2 |
| Proportion of female hospitalized by age group, % | 42,4 | 47,2 | 53,9 | 65,2 | 72,8 |

| **Lethality** |  |  |  |  |  |
| --- | --- | --- | --- | --- | --- |
| Number of deaths among those hospitalized, abs. | 15 | 35 | 141 | 282 | 65 |
| Proportion of age group among all deaths, % | 2,8 | 6,5 | 26,2 | 52,4 | 12,1 |
| Proportion of deaths in the age group, % | 2,4 | 3,1 | 6,9 | 17,0 | 38,5 |
| Number of male deaths by age group, abs. | 7 | 26 | 85 | 115 | 14 |
| Number of female deaths by age group, abs. | 8 | 9 | 56 | 167 | 51 |
| Proportion of male deaths by age group, % | 46,7 | 74,3 | 60,3 | 40,8 | 21,5 |
| Proportion of female deaths by age group, % | 53,3 | 25,7 | 39,7 | 59,2 | 78,5 |

\* the longevity age group ("91+") includes age cohorts from 91 to 100 years.

For a more detailed study of those hospitalized with COVID-19 and mortality among them, the study sample, in addition to the WHO classification, was divided into 83 age cohorts (in the context of this article, one age cohort consists of people of the same birth year), of which 42 are of working age (18-59 years) and 41 are of retirement age (60-100 years).

### Hospitalized, baseline data

The distribution of those hospitalized in absolute numbers by age in the original data is shown in **Figure 1**.

**Figure 1.**
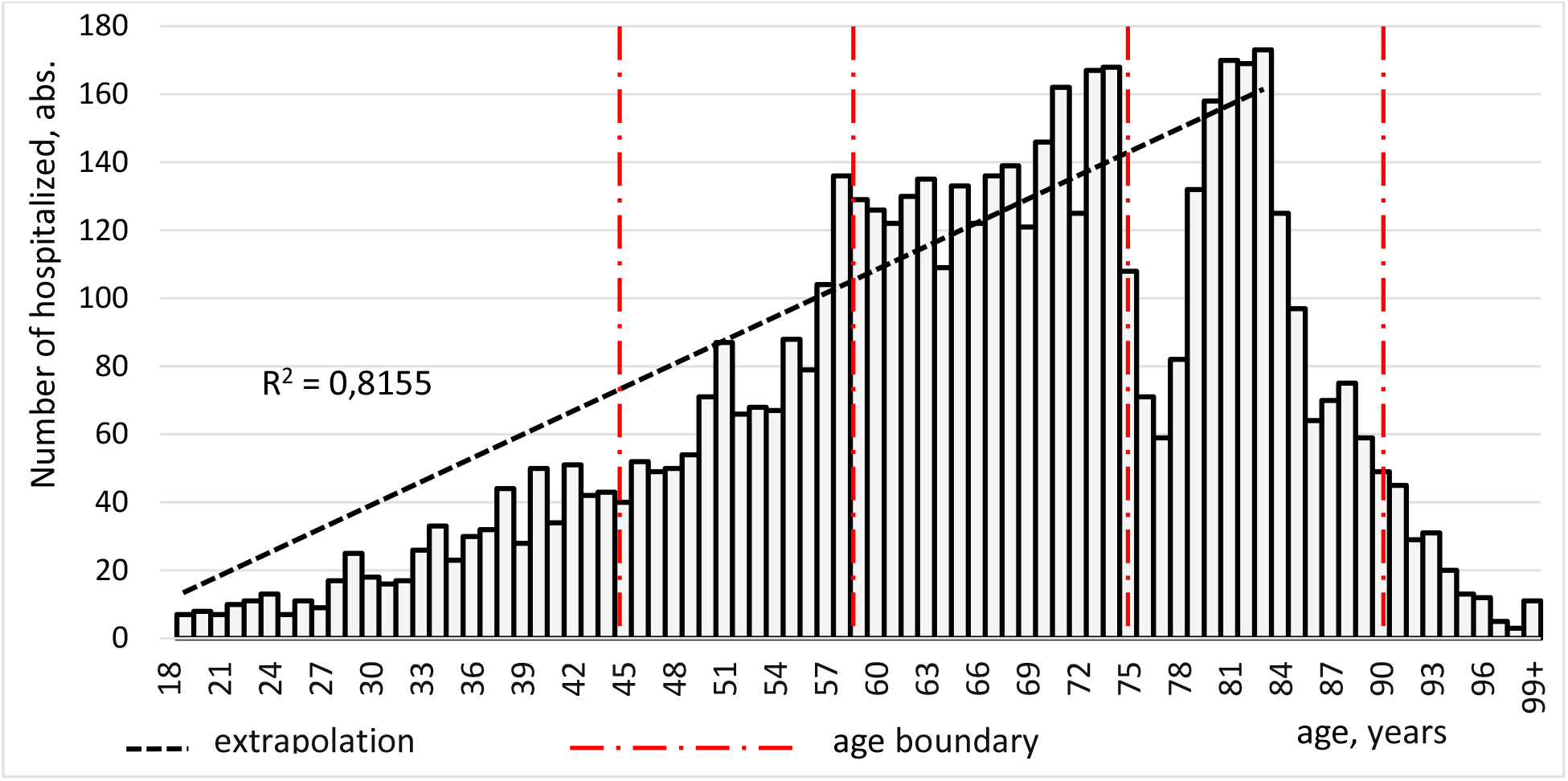
Distribution of hospitalized with COVID-19 by age in the original data, abs.

As can be seen from figure 1, the young and middle-aged groups do not show a normal distribution, with a mean age closer to the right border of the age group (39.0 ± 5.0 and 53.1 ± 4.3, respectively).

The absolute number of hospitalizations with COVID-19 rises from a minimum at 18 years of age to a maximum at the age of 83. The extrapolation trend line for the increase in hospitalizations is a straight line with a coefficient of determination (goodness of fit) of R^2^ = 0.8155. Between the ages of 84 and 100, the number of hospitalizations decreases sharply. The observed sharp decrease in the absolute number of those aged 75-79 years is not due to any age or physiological characteristics, but to a demographic failure in the birth rate between 1942 and 1946 [30]. The small number of those hospitalized at ages 90+ (only 169, or 3.0%) is proportional to the proportion of these ages in the population structure of Moscow, which is about 2.7% [30].

The distribution of hospitalized persons by age group in the proportions in the raw data is shown in **Figure 2A**.

**Figure 2.**
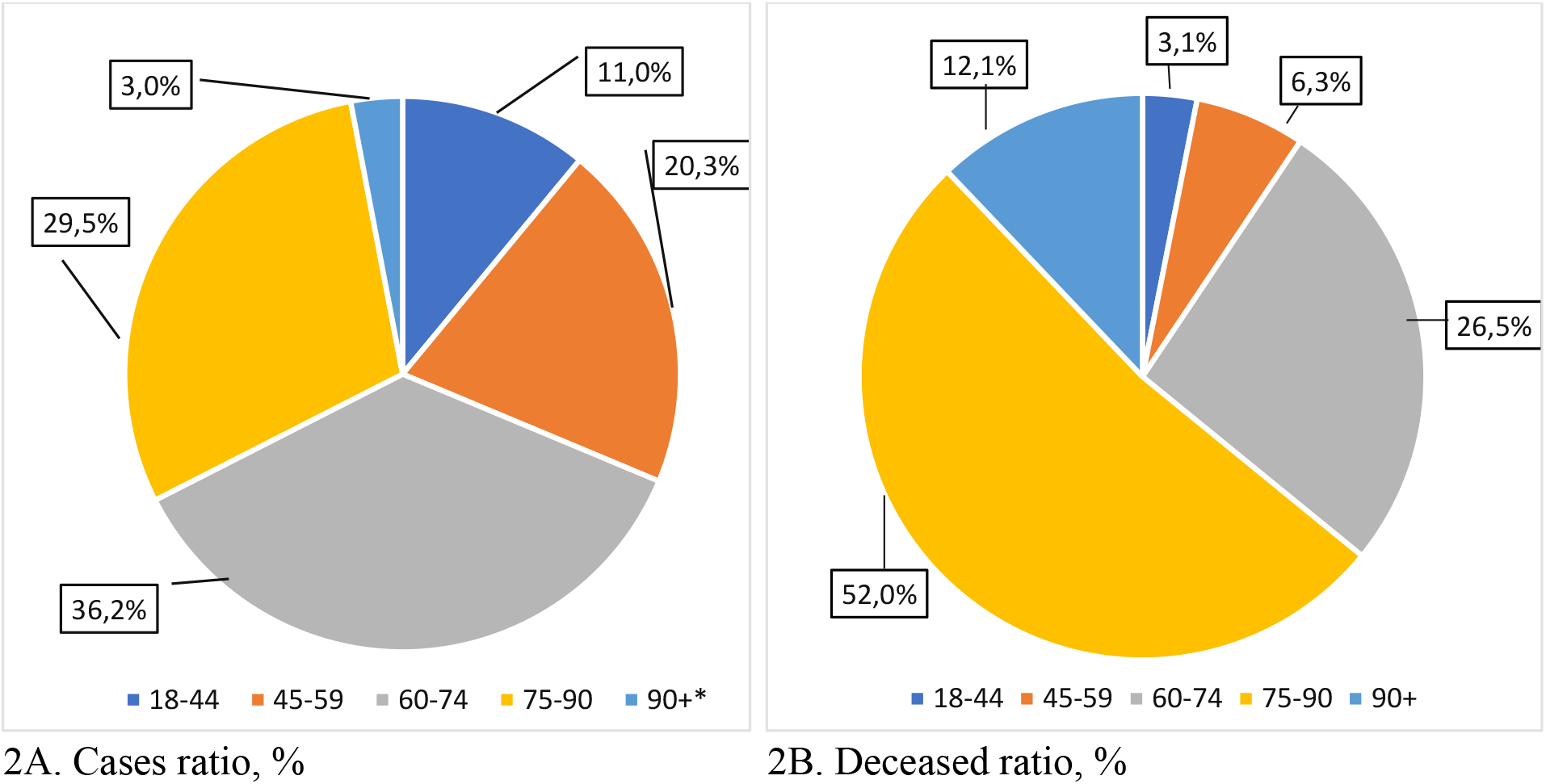
Distribution of hospitalized (A) and deceased (B) by age group in the initial data, %.

The group with the highest absolute number of hospitalizations is the elderly - 2041 people or 36.2%. A total of 3 of the 5 groups (middle age, old age and old age) determine 86.0% of the total number of all hospitalized, representing 4,842 out of 5,629 people. The younger age group accounts for 618 persons (11.0%). The smallest number of hospitalized persons is registered in the “longevity” group - 169 persons or 3.0%.

The distribution of hospital admissions by age, shown in Figures 1 and 2, clearly shows that the risk groups for the absolute number of cases are «45-90 years old».

When analyzing baseline data on the gender proportions (men and women) of those hospitalized with COVID-19 according to age (**Fig. 3A**), it follows that the main risk group is women in the ‘old age’ (75-90 years) and ‘longevity’ (90+ years) groups, who are hospitalized at approximately 1.9 to 2.7 times the risk of men (65.2% and 72.8%, respectively). A less pronounced but significant risk group is young men (18-44 years). When admitted to hospital with COVID-19, they require inpatient care about 1.4 times more often than women of the same age. Men and women in the ‘middle-aged’ and ‘elderly’ groups (45-59 and 60-74 years, respectively) are hospitalized at about the same rate of about 50%.

**Figure 3.**
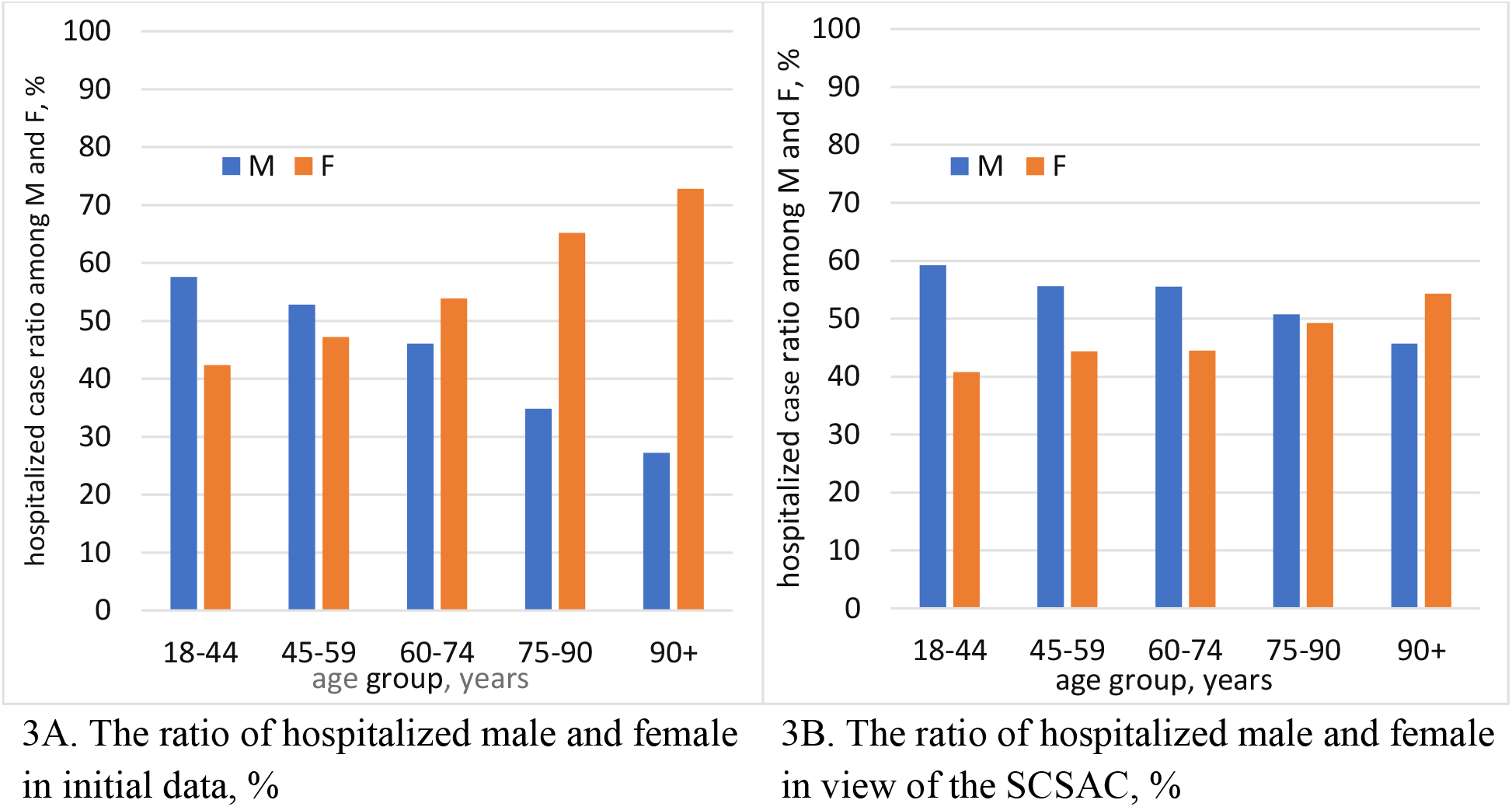
Proportions of male (M) and female (F) hospitalized with COVID-19 in baseline (A) and estimated data including SCSAC (B), %.

Summarizing the results of the analysis of baseline hospitalization data (Figures 1, 2A, 3A), it can be concluded that the main risk groups for hospitalization in terms of gender and age are women in the elderly and old age groups. The total number of hospitalized women aged 60-90 years during the period under review was 1,183 out of 5,629 hospitalized, or 38.8%.

### Lethality, baseline data

The distribution of the absolute number of deaths by age cohort in the original data is shown in **Figure 4**. The extrapolation curve of the increase in the absolute number of deaths from 18 to 83 years is described by a third-degree polynomial with a high coefficient of determination (R^2^ = 0.8111). At ages 18 to 56 years, 19 of the 39 age cohorts reported no fatalities among those hospitalized with COVID-19 over the entire analysis, while the remaining 20 cohorts reported only a single fatality. The proportion of fatal cases for ages 18-56 years (in fact, the main working ages) is approximately 2.7%.

**Figure 4.**
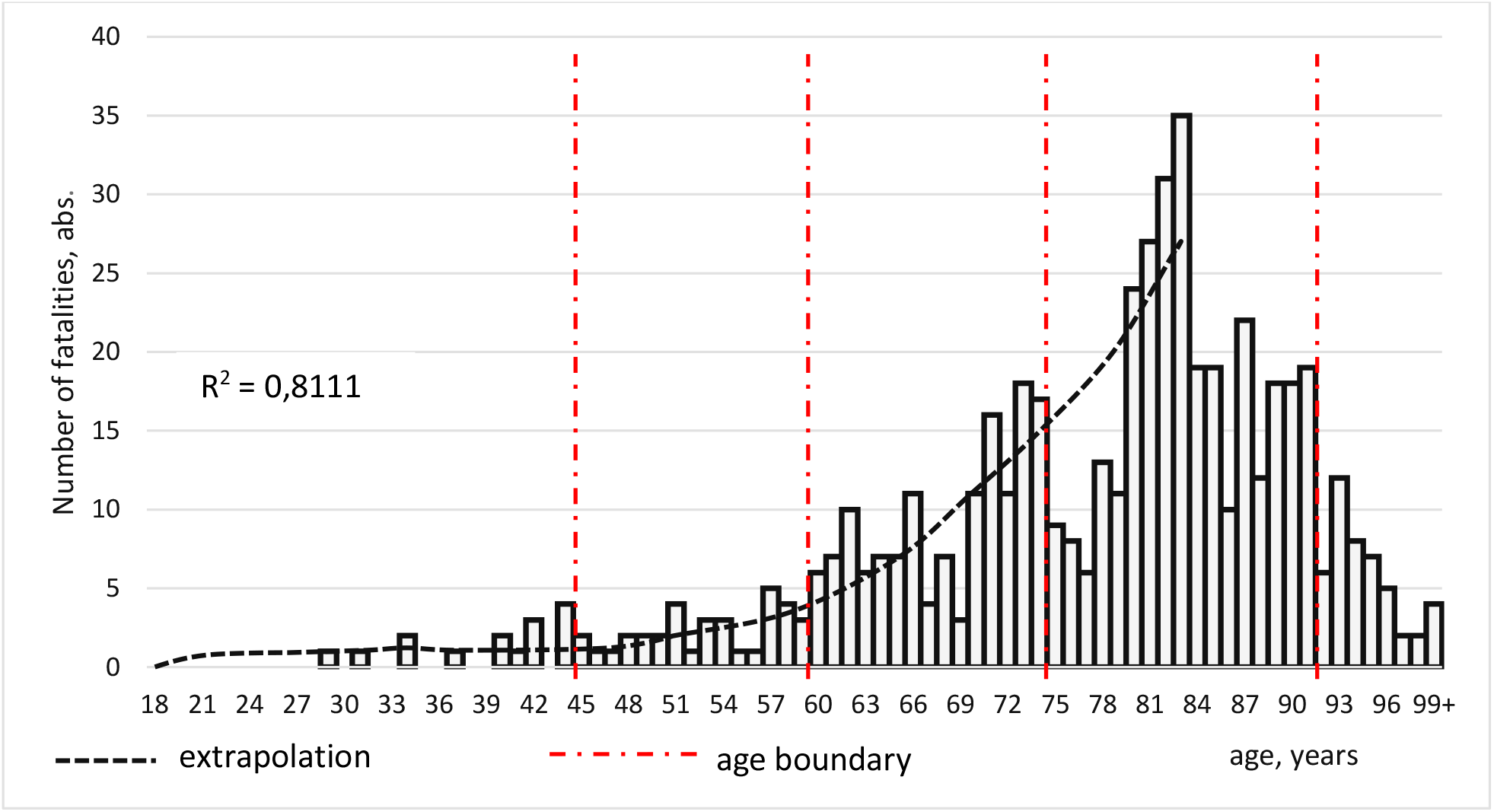
Distribution of COVID-19 deaths by age cohort, abs.

The rise in the absolute number of deaths begins at age 57 (almost the beginning of the retirement ages) and reaches a maximum at age 83. The sharp decline in the absolute number of deaths at ages 75-79 - as in the case of the decline in the absolute number of cases hospitalized with COVID-19 at these ages - is explained, we think, by the collapse in fertility between 1942 and 1946. [30]. By age 99+, the absolute number of deaths fell from 18-19 to 2 per age cohort. When estimating the decline in absolute numbers of deaths at ages above 83 years, the proportion of cases in the 91+ age group of all cases must not exceed 3.0%.

Comparative analysis of the growth curves of the absolute number of hospitalizations with COVID-19 (Fig. 1) and deaths among them (Fig. 4) as a function of age shows that in both cases the maximum values are reached by the age of 83. The extrapolation curves for both hospitalization and mortality at ages 18-83 have high and very close coefficient of determination values (0.8155 and 0.8111, respectively). But the extrapolation curves themselves are described by fundamentally different formulas: a straight line for hospitalization and a third-degree polynomial for lethality.

The proportion of COVID-19 fatalities for each age group in the normalized rates (%) is shown in **Figure 5**.

**Figure 5.**
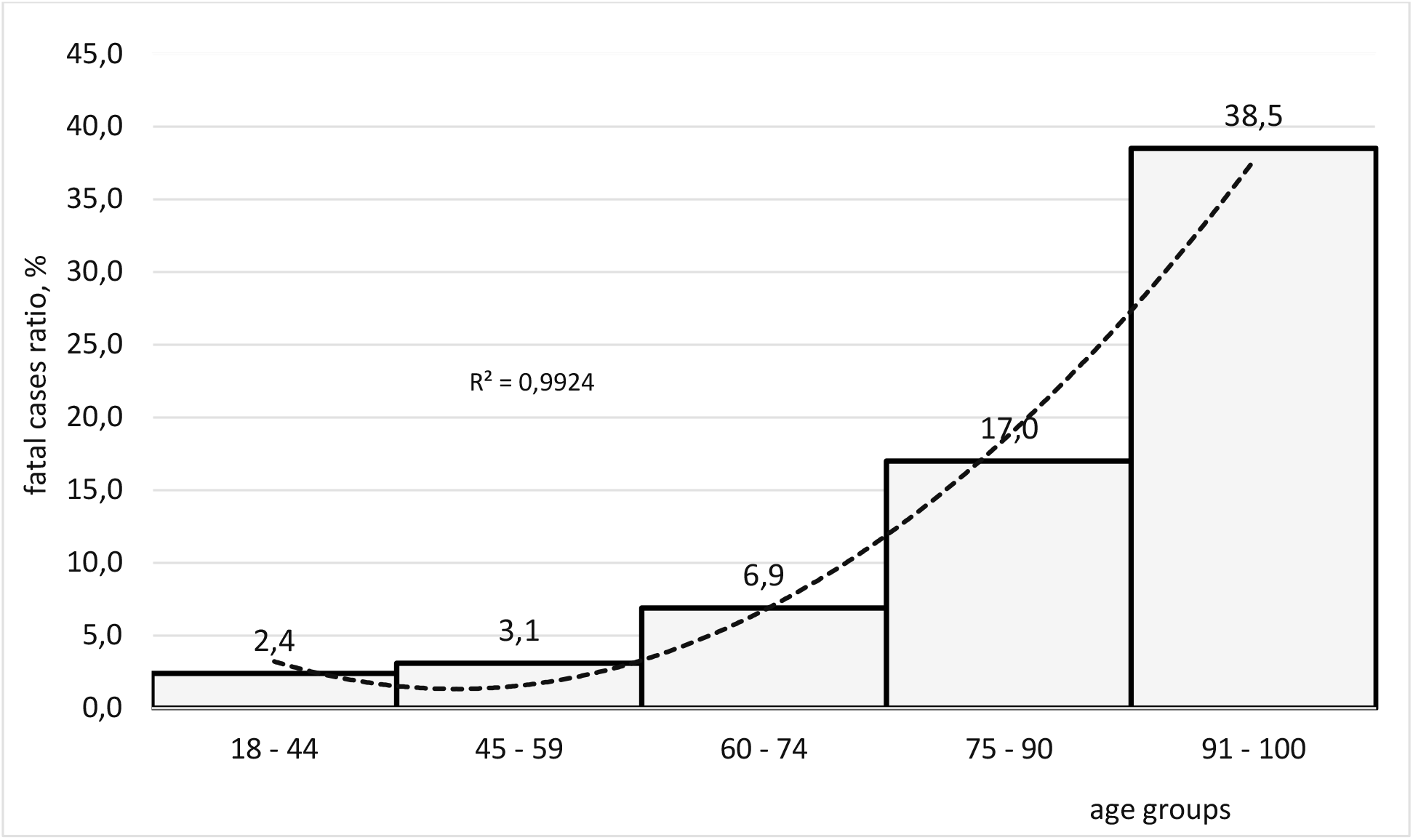
The ratio of deaths from COVID-19 in age groups, %.

In the normalized figures, the maximum number of deaths, unlike the absolute numbers, shifts from the age of 83 to the “91 to 100 years” group and, based on the results of the absolute and relative mortality analysis, the two age groups (75 to 90 and 91 to 100 years) are the main risk groups for deaths, accounting for 55.5 per cent of all deaths.

The extrapolation curve “mortality as a function of age” (Fig. 5) is a quadratic function with a very high approximation reliability (R^2^ = 0.9924), indicating a strong association between patient age and the possibility of death for patients. The conclusion about a more severe course and more likely lethal outcome with increasing patient age is rather trivial, but the resulting curve has numerical properties and can be of practical value and used as a rough prognostic guide in assessing the course and outcome of COVID-19 in health care institutions. Of course, the findings need to be independently verified and validated in relevant samples for those hospitalized in other hospitals in Moscow or other regions of Russia (or countries of the world).

The different types (and formulas) of extrapolation curves - a straight line for hospitalization (Fig. 1) and a polynomial of degree 2 or 3 for mortality (Figs. 4 and 5) - indicate, in our opinion, that in the studied ages 18-100 years, mortality is not only determined by age, but also by additional factors whose identification and analysis requires an independent investigation.

The absolute distribution in the raw data of deaths among the hospitalized in all age groups studied is shown in the proportions in Figure 2B.

Comparing Fig. 2, charts A (hospitalization rate, %) and B (death rate, %) shows that 42 cohorts at ages 18-59 years, constituting the main working age population in Moscow (and Russian Federation), account for 31% of all hospitalizations, and 41 retirement age cohorts, 69%. As for fatal outcomes, the working-age cohorts account for only 9% of all deaths and the retirement-age cohorts for 91% (**Table 2**). That is, the same number of age cohorts (42 and 41) in the working-age and retirement-age categories yield a ratio of “1 : 2.2” (31 / 69%) for hospitalizations and “1 : 10” for deaths (9 / 91%).

**Table 2.** The proportion of hospitalization and mortality in cohorts of working and retirement age.

| Age cohorts | Number of age cohorts, abs | Hospitalization, % | Mortality, % |
| --- | --- | --- | --- |
| Working ages, 18 – 59 years old | 42 | 31,3 | 9,3 |
| Retirement ages, 60 – 100 years old | 41 | 68,7 | 90,7 |

The results of the proportion in the normalized rates (%) of male and female deaths in each age group according to the baseline data are presented in **Figure 6 A**.

**Figure 6.**
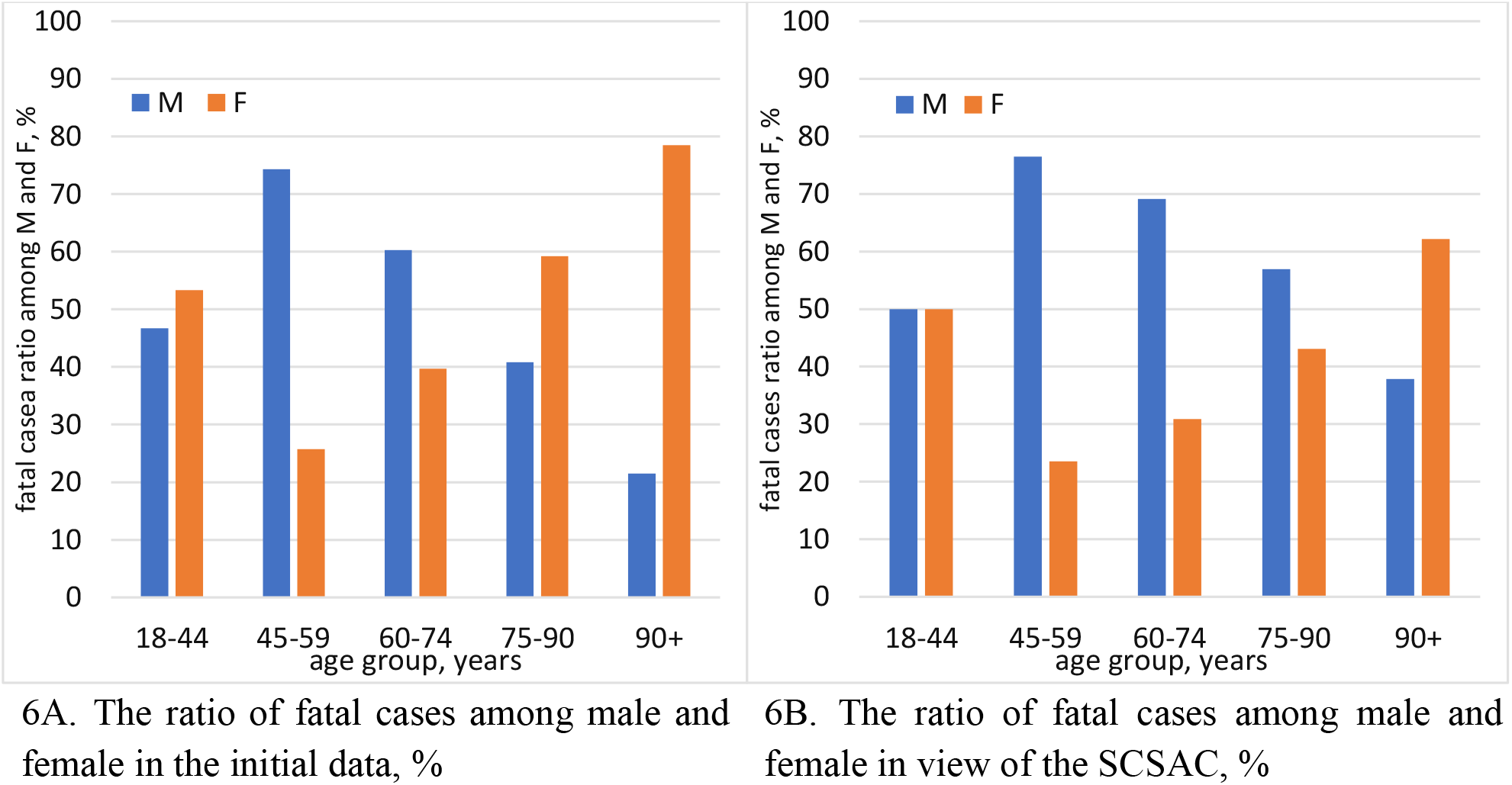
The ratio of fatal cases in COVID-19 among male (M) and female (F) in initial (A) and calculated data in view of the SCSAC (B), %.

As can be seen in Fig. 6A, at “young ages” (18-44 years) the mortality rate among men and women differs very little and is in the region of 50% (46.3 vs. 53.3%). Between the ages of 45 and 100, the reported relative mortality among men and women is almost a mirror image. While in men the proportion of fatal cases shows a maximum value of 74.3% in the group of 45-49 years old and by 100 years old it decreases linearly to 22%, among female patients we observe a diametrically opposite picture - a linear increase from 25.7% to 78.5%. In the “45-59” and “60-74” groups, the proportion of men who died is higher than that of women by about 3 and 1.5 times, respectively. In the age groups “75-90” and “91+” the situation is reversed and the proportion of female deaths prevails over male deaths: in these ages women die about 1.5 and 3.7 times more often than men. The use of Chi-square test suggests that there is a statistically significant relationship between age and sex for mortality (p-value <0.05), but the relationship is rather weak - Cramer’s coefficient is 0.27.

Thus, the analysis in the normalized baseline data of the dependence of mortality in COVID-19 on gender and age characteristics (Fig. 6A) does not clearly identify the main high-risk group. The maximum-risk groups are equally middle-aged men and longevity women, and the high-risk group is elderly men and senile women.

Summarizing the above epidemiological analysis of the baseline data on patients hospitalized with COVID-19, the main risk groups for both hospitalization and mortality according to gender and age can be identified as follows: the highest risk groups for hospitalization are elderly and senile women, and for mortality are men aged 45 to 59 and women aged 91 to 100.

The results of the analysis of hospitalization and mortality in COVID-19, both in absolute values and in normalized figures, have important scientific and practical significance in the planning of medical care for hospitalized patients, taking into account their age (bed stock, priority set of drugs, etc.). These results can also be used in studies to assess the economic costs of COVID-19, including both direct costs of hospitalization and indirect costs in the case of fatalities in patients of working age.

However, in our opinion, these conclusions are true provided that the number of men and women in all age cohorts in Moscow is equal, i.e. their proportion does not change depending on age and is always “1 : 1”. The real situation is that this proportion does not hold in any of the age groups we studied. With increasing age, the number of men in the population decreases much faster than the number of women. According to data published by Rosstat on the population by sex and age in Moscow as of January 1, 2020, the number of men in the population becomes smaller than the number of women by a factor of 1.1 (for ages “18-45”) to 2.3 (for ages “90+ “) [30].

To exclude the influence of this disproportion on the conclusions about the true risk groups for hospitalization and mortality, we performed statistical correction of the sex and age composition (SCSAC) of sick people in the studied age groups. The approach in using the SCSAC is similar in principle to that of rationing the incidence rate per 100,000 population. The SCSAC coefficients obtained on the basis of Rosstat data for each age group made it possible to achieve the condition that the number of men and women in each age group would be the same. This approach to analyzing morbidity and mortality makes it possible to calculate the incidence of diseases and deaths from COVID-19, excluding the influence of gender imbalances in the age groups under study and formulating more substantiated conclusions.

The number of cases and deaths by sex and age are distributed differently when using the SCSAC than when analyzing the absolute baseline data. Processing the baseline data using the SCSAC coefficients to calculate the proportions of males and females in the age groups leads to adjustments or changes in the definition of the main risk groups by sex and age for both morbidity and mortality.

The dimensionless coefficients of the SCSAC we calculated for each age group increase from 1.1 for the ‘young age’ group to 2.3 for the ‘longevity’ group.

### Hospitalized, data including SCSAC

The proportion of male and female hospitalized in each age group, taking into account the SCCVS, is shown in Figure 3B.

The data in **Table 3** and Fig. 3B shows that the proportion of hospitalized males decreases from 60 to 46 per cent as the age rises from 18 to 100. This trend can be seen both in the proportion of males and females hospitalized in the original data and in the statistical distribution of males and females in the general population in Moscow. On the basis of the chi-square test, we can conclude that there is a statistically significant association between age and gender among the COVID-19 cases (p-value < 0.05). However, the Cramer’s coefficient V as a measure of the strength of the association shows that the association is rather weak (V=0.16), indicating that in addition to gender and age, other factors influence hospitalization for COVID-19 disease, such as the presence of comorbidities, their number, presence or absence of vaccination, individual immune system status, etc..

**Table 3.** Distribution of hospitalized with COVID-19 by gender and age (years) groups, taking into account the SCSAC.

| Age group | Young age | Middle age | Old age | Senile age | Longevity |
| --- | --- | --- | --- | --- | --- |
| Age cohorts, year old | 18-44 | 45-59 | 60-74 | 75-90 | 90+ |
| **Hospitalized, initial data** |  |  |  |  |  |
| The number of hospitalized males, abs. | 356 | 602 | 941 | 578 | 46 |
| The number of hospitalized females, abs. | 262 | 538 | 1100 | 1083 | 123 |
| Percentage of male, Rosstat [binomial confidence interval], % | 48 [53, 61] | 47 [50, 55] | 41[44, 48] | 34 [32, 37] | 31 [21, 34] |
| **Hospitalized, calculated data in view of the SCSAC** |  |  |  |  |  |
| SCSAC coefficient number of females per 1 male, Rosstat | 1,07 | 1,12 | 1,46 | 1,93 | 2,25 |
| The number of hospitalized males, abs. | 356 | 602 | 941 | 578 | 46 |
| The number of hospitalized females, abs. | 245 | 480 | 753 | 561 | 55 |
| Total number of hospitalized (male and female), abs. | 601 | 1082 | 1694 | 1139 | 101 |
| Percentage of hospitalized male in the age group, % | 59,2 | 55,6 | 55,5 | 50,7 | 45,7 |
| Percentage of hospitalized female in the age group, % | 40,8 | 44,4 | 44,5 | 49,3 | 54,3 |

From the raw data (Figure 3A), the proportion of hospitalized women in the age categories “60-100 years” increases compared to the proportion of hospitalized men in the corresponding age categories and, as mentioned in our work above, it seems that women in the elderly and old age categories constitute the main risk group for hospitalization. The binomial criterion showed that in the age cohorts “18-74”, the age and sex composition of hospitalized people differed significantly from the age and sex composition of the Moscow population as a whole.

When the study groups were rationed using the RASC (Figure 3B), the group at highest risk for hospitalization with COVID-19 was young men “18-44 years” (mean age 39.0 ± 5.0 years), who were hospitalized almost 1.5-fold more often than women. The relative risk group are middle-aged and elderly men (“5-59” and “60-74” respectively). They are hospitalized 1.3 times more often than women in the corresponding age groups. In the ‘old age’ group, more men than women are hospitalized (56 versus 44%), whereas the reverse was true at the start, with 46% of men versus 54% of women hospitalized. The distribution of proportions of hospitalized males and females in the “75–90” age group changed fundamentally from 35% and 65% at baseline to 51% and 49% with the use of the RHMS. The proportion of men and women hospitalized in the “91-100” age group also changed significantly: the proportion of women remained higher than that of men, but decreased from 73% to 54%, a decrease of almost 1.4-fold.

Thus, in 3 of the 5 age groups (or 57 of the 83 age cohorts) the rate of hospitalization among men is higher than among women. The true high-risk group for hospitalization with COVID-19 is men aged “18-44” (mean age 39.0 ± 5.0 years). Also significant risk groups are middle-aged and elderly men. That is, the main cohort for hospital admissions with COVID-19 are men of prime working age.

Social and physiological characteristics of men contribute to the higher rate of hospitalization among male compared to female.

Social factors include social activity, working age, increased mobility, and not always responsible attitudes towards anti-epidemic measures, in particular mask compliance. Our findings are confirmed by other researchers who attribute the high risk of COVID-19 in young men to higher rates of smoking, lower hygiene (hand washing), prior respiratory disease [35].

Physiological features in young men T. Osama et al. attribute this to the presence of elevated levels of sACE2 in their plasma. The angiotensin-converting enzyme receptor (sACE2) is the receptor required for SARS-CoV-2 to enter the target cell. Serum sACE2 levels have also been found to correlate strongly with immune receptors in the lungs of men and the elderly and to be less correlated in women and children [36]. Several studies have observed that sACE2 tends to increase with age and that men showed higher plasma sACE2 concentrations compared with children and women [37, 38].

For the reasons indicated, young men (18-44 years of age) maintain an active epidemic process, representing numerous sources of infection for other categories of citizens. They pose a particular risk for the elderly and the elderly (age cohorts “60-74” and “75-90”), who are the main contributors to mortality rates from COVID-19.

### Lethality, data including SCSAC

Of the sample analyzed (5629 COVID-19 cases), the number of deaths was 538, of whom 247 were men and 291 women. The distribution of deaths by gender and age according to baseline data is shown in Table 1 and Fig. 6?.

Estimated data for an age and sex-specific analysis of mortality with regard to SCSAC are presented in **Table 4**.

**Table 4.** Distribution of the deceased due to COVID-19 by sex and age characteristics in view of the SCSAC.

| Age group | Young age | Middle age | Old age | Senile age | Longevity |
| --- | --- | --- | --- | --- | --- |
| Age cohorts, year old | 18-44 | 45-59 | 60-74 | 75-90 | 90+ |
| **Mortality, initial data** |  |  |  |  |  |
| The number of deaths, abs | 15 | 35 | 141 | 282 | 65 |
| The number of deaths male, abs | 7 | 26 | 85 | 115 | 14 |
| The number of deaths female, abs | 8 | 9 | 56 | 167 | 51 |
| **Mortality, calculated data in view of the SCSAC** |  |  |  |  |  |
| SCSAC coefficient number of females per 1 male, Rosstat | 1,07 | 1,12 | 1,46 | 1,93 | 2,25 |
| The number of deaths male, abs | 7 | 26 | 85 | 115 | 14 |
| The number of deaths female, abs | 7 | 8 | 38 | 87 | 23 |
| Total number of deaths (male and female), abs | 14 | 34 | 123 | 202 | 37 |
| Proportion of deceased male, % | 50,0 | 76,5 | 69,1 | 56,9 | 37,8 |
| Proportion of deceased male female, % | 50,0 | 23,5 | 30,9 | 43,1 | 62,2 |

The percentage distribution of deaths for males and females in each age group, taking into account the SCSAC, is shown in Figure 6B.

An analysis of mortality using the SCSAC (Figure 6B) shows that for young and middle-aged patients (“18-44” and “45-59” years old), the “male : female” ratio in relation to baseline data is almost unchanged, remaining approximately “1 : 1” (young) and “3 : 1” (middle-aged). In the older age group (60-74 years) the male fatality rate increases from 60% to 69% and the “male : female” fatality rate changes from around “1.5 : 1” at baseline to around “2.3 : 1” for the SCSAC option. That is, the risk of death for males in this age group increases by a factor of approximately 1.5 when compared to the estimated risk for males based on the baseline data. In the older age group, a fundamental redistribution of the gender proportions of fatalities was registered: the proportion of fatal cases in men became higher than in women and rose from 41% to 57%. The proportions in the longevity group have also changed significantly. Lethality among women - as in the original data - remained higher than among men. However, while women were about 3.6 times more likely to die than men in the baseline data, the mortality rate among women is only 1.7 times higher than among men when converted using the SCSAC.

In 3 out of 5 age groups (46 out of 83 age cohorts), the proportion of fatal cases among men is higher than among women. When calculations are adjusted for COVID-19, the true group with the highest risk of death from COVID-19 is middle-aged and elderly men (mean age 53.1 ± 4.3 and 68.3 ± 4.5 years), for whom the probability of death is 3.3 and 2.2 times higher than for women of the same age, respectively. Men of advanced age are also at increased risk of mortality.

Thus, the true maximum risk group for hospitalization with COVID-19 is young men. Middle-aged and elderly men can be classified as being at increased risk. The risk of mortality is defined as maximum among middle-aged and elderly men, and increased among older men.

## Conclusion

Our study identified a number of important gender- and age-specific characteristics and patterns of the COVID-19 epidemic among hospitalized adults and identified gender- and age-specific risk groups who are most at risk of hospitalization and the potential for death from COVID-19 disease.

We were the first in the world to substantiate the feasibility of using statistical sex-age adjustment (SAVA) to equalize existing population disparities between males and females of the same age category in the administrative territory under study. Our proposed rationing approach establishes the true risk groups for hospitalization and mortality in COVID-19 disease.

The results obtained at baseline do not support the conclusions of other researchers that the risk groups are elderly men. As described above, our results suggest that the risk groups for hospital admissions are women (elderly and seniors) and that both men (middle-aged group) and women (longevity group) are at risk for mortality. It is possible that these differences in risk group estimates are due to age and sex differences between populations around the world, due to, for example, different life expectancy of men or other reasons, but their clarification was not the aim of our work and could be an independent research.

It is important to note that by using the approach of statistical correction of the sex and age composition of the Moscow population, we independently confirmed the existing estimates of a number of researchers from different countries of the world that the main risk group for possible lethal outcomes is males. In addition to the pre-existing results, our study has expanded knowledge about the age structure of men as a risk group for fatal outcomes: the risk groups are not only elderly men, but also middle-aged and elderly men.

We proved a statistically significant association of hospitalization and mortality from COVID-19 with gender and age (p-value <0.05). The strength of the association (Cramer’s measure) was weak (V = 0.16 and 0.27, respectively), suggesting that hospitalization and mortality from COVID-19 in the Moscow population are influenced by additional factors (other than gender and age), which require separate investigation.

We also found and substantiated that young men (mean age 39.0 ± 5.0 years) are the group at highest risk of hospitalization for COVID-19 disease. They are 1.5 times more likely to be hospitalized with COVID-19 than young women.

In terms of COVID-19 deaths, middle-aged and elderly men are the main risk group. For them, the chances of dying when hospitalized with COVID-19 are about 3.3 and 2.2 times higher than for women of the same age.

These patterns suggest that increased attention should be paid to identified gender- and age-specific categories in the design of counter-epidemic measures and in the provision of inpatient health care.

## Data Availability

All the data obtained in this study are available to the authors upon reasonable request

https://rosstat.gov.ru/storage/mediabank/Bul_chislen_nasel-pv_01-01-2021.pdf

## Acknowledgement

Authors express their gratitude to Andrey N. Gerasimov, PhD (Physics and Mathematics), leading research fellow in the Central Research Institute of Epidemiology, Federal Service for Surveillance on Customer Rights Protection and Human Wellbeing; Olga A. Manerova, PhD, MD, professor of the Department of Public Health and Healthcare n.a. N.A. Semashko First Sechenov Moscow State Medical University (Sechenov University); Anna D. Muzyka, senior research fellow in the Central Research Institute of Epidemiology, Federal Service for Surveillance on Customer Rights Protection and Human Wellbeing; employees in the Central Research Institute of Epidemiology of Rospotrebnadzor Yulia R. Melnichenko and Anton A. Privalenko for his help in data collection, editing, discussion and valuable critical comments while working on this article.

